# Transcranial alternating current stimulation for Parkinson’s disease: a systematic review and meta-analysis of motor outcomes

**DOI:** 10.64898/2026.08.21.26360996

**Authors:** Thuy Tien Mai, Tea Gjishti, Karsten Witt, Mandy Roheger, Christoph S. Herrmann

**Author notes:** Contributing authors.

## Abstract

Transcranial alternating current stimulation (tACS) is a promising noninvasive intervention for modulating pathological brain oscillations in Parkinson’s disease (PD). To evaluate its clinical and neurophysiological efficacy, we searched five databases (Web of Science, PubMed, Scopus, Google Scholar and APA PsycInfo) up to August 31, 2025, for trials employing tACS in patients with idiopathic PD. Risk of bias was assessed using the RoB 2 and ROBINS-I tools. Random-effects meta-analyses were used to calculate standardized (SMD) and unstandardized mean differences (MD) with 95% confidence intervals (CIs). We included 10 studies (184 patients with PD, mean age: 64.9, mean disease duration: 5.2 years) in the qualitative review and seven trials (146 patients with PD, mean age: 65.6, mean disease duration: 5 years) in the meta-analysis. No statistically significant differences favoring active tACS over control were found in overall motor severity (UPDRS: SMD = 0.21, 95% CI [–0.10, 0.52], ***p* = 0.097**), tremor (SMD = –0.40, 95% CI [–1.97, 1.17], ***p* = 0.478**), or a neurophysiological marker of inhibitory response, represented by short intracortical inhibition (MD = 0.00, 95% CI [–0.40, 0.41], ***p* = 0.971**). The prediction intervals indicated substantial uncertainty, and significant between-study heterogeneity was observed, particularly for tremor outcomes (***I***^2^ **= 86.1%**). This variability and limitation in evidence quality is largely driven by small sample sizes, highly heterogeneous stimulation protocols, and varying outcome assessments. Systematically, tACS was generally well-tolerated, with no serious adverse events reported across the included studies; however, formal safety assessment was beyond the scope of this review. Current exploratory evidence shows a lack of consistent improvements in motor symptoms or functions in PD largely due to protocol-level heterogeneity. Future studies should consistently assess the MDS-UPDRS III post-tACS and report its specific subscores alongside neurophysiological measures to enable robust meta-analyses.

## 1 Introduction

Parkinson’s disease (PD) is a progressive neurodegenerative disorder clinically characterized by cardinal motor symptoms, including bradykinesia, resting tremor, and rigidity ^1^. Pathophysiologically, these motor deficits are tightly linked to the degeneration of dopaminergic neurons in the substantia nigra, leading to aberrant neural oscillatory activity within the cortico-basal ganglia-thalamocortical network. Specifically, pathological synchronization in the beta frequency band (13–30 Hz) is recognized as a neurophysiological hallmark of bradykinesia and rigidity ^2,3^, while distinct oscillatory networks, often involving the cerebellum and primary motor cortex (M1), are implicated in tremor generation ^4^. Importantly, beyond pathological synchronization, impaired cortical compensatory recruitment is increasingly recognized as a relevant component of PD motor dysfunction ^5,6^, suggesting that both subcortical and cortical mechanisms should be considered in therapeutic targeting.

Due to these aberrant network dynamics, PD is increasingly conceptualized as an “oscillopathy”^3^. This underlying oscillatory dysfunction, in turn, drives measurable alterations in neurophysiological markers, such as corticospinal excitability and initracortical inhibition. Research has demonstrated that motor evoked potentials (MEPs) generated by single-pulse (SP) transcranial magnetic stimulation (TMS) show impaired motor recruitment in PD ^7^. Conversely, intracortical inhibitory systems involving Gamma-Aminobutyric acid A (GABA_A_)-mediated pathways—which can be assessed through the paired-pulse (PP) TMS protocol of short-interval intracortical inhibition (SICI) applying a conditioning stimulus at a subthreshold intensity followed by a test stimulus at suprathreshold intensity after 2 ms—tend to be reduced, reflecting deficient GABA_A_ activation ^7^.

While pharmacological dopamine replacement remains the therapeutic gold standard, its long-term utility is frequently limited by fluctuating efficacy and the emergence of debilitating side effects ^8,9^. Furthermore, although deep brain stimulation (DBS) represents a highly effective invasive neuromodulation strategy, its application is constrained by stringent surgical eligibility criteria and potential procedural risks ^10^. Consequently, there is a growing interest in non-invasive brain stimulation (NIBS) techniques as adjunct therapies. Among these, transcranial alternating current stimulation (tACS) has emerged as a particularly promising tool ^11^. By delivering weak alternating electrical currents to the scalp, tACS can exogenously entrain or disrupt targeted endogenous cortical oscillations ^12^. In the context of PD, this provides a compelling mechanistic rationale: utilizing frequency-specific tACS to non-invasively modulate the pathological brain rhythms driving motor impairment. However, conventional scalp tACS is expected to influence superficial cortical circuits more directly than deep subcortical nuclei. Therefore, in PD, tACS may be better conceptualized as a tool to modulate cortical network dynamics (including compensatory mechanisms) rather than as a direct analogue of DBS for subcortical target engagement.

Because tACS is intended to modulate oscillatory network dynamics, neurophysiological end- points are not merely ancillary outcomes but mechanistically informative measures. In particular, TMS-derived markers such as SICI can quantify cortical inhibitory dysfunction, probe target engagement, and potentially help identify response profiles to stimulation. Thus, tACS and TMS-based neurophysiology can be viewed as complementary approaches: the former aims to modulate dysfunctional network activity, whereas the latter characterizes cortical excitability and treatment-related physiological change ^13^.

Despite the strong theoretical and neurophysiological rationale for tACS in PD, the literature remains highly fragmented. Current tACS literature in PD is characterized by profound methodological heterogeneity. Studies vary drastically in stimulation parameters (frequency, intensity, montage), patient characteristics, and chosen endpoints. This lack of standardization and outcome consistency makes it difficult to ascertain the true therapeutic potential of tACS for patients with PD.

To date, broader systematic reviews of NIBS have predominantly evaluated transcranial direct current stimulation (tDCS) and repetitive transcranial magnetic stimulation (rTMS) for PD, often leaving tACS only briefly addressed ^14–19^. Consequently, a comprehensive quantitative synthesis evaluating the dual clinical and neurophysiological impact of tACS remains a critical gap in the literature.

While a recent meta-analysis by Ye et al. (2025) ^20^ has contributed to the understanding of neurophysiological tACS effects in PD, their analytical approach—which incorporated healthy control comparators alongside clinical samples—may obscure disease-specific effects. Specifically, pooling PD patients with healthy controls conflates therapeutic efficacy (changes within PD patients) with mechanistic differences between groups, potentially diluting the signal of clinically meaningful improvements and preventing clear assessment of whether tACS produces differential benefits in PD populations. Furthermore, divergent data extraction and synthesis methods in that work have yielded effect size estimates that contrast with some primary study reports. These differences in methodological perspective underscore the utility of a further, strictly clinical synthesis to provide a highly transparent and reproducible estimate of tACS efficacy.

Therefore, the primary objective of this systematic review and meta-analysis is a rigorous evaluation of the current evidence regarding tACS efficacy in PD. To resolve existing literature inconsistencies, we strictly compare active tACS against sham or control conditions within clinical PD populations. This approach encompasses studies evaluating both single and multi-session protocols, regardless of the stimulation frequency, intensity or location applied.

Our analysis evaluates the clinical utility of tACS across two primary domains: (1) **clinical motor symptoms**, representing observable behavioral changes such as overall motor severity and tremor; and (2) **neurophysiological motor function**, focusing on markers of inhibitory response, specifically TMS-derived SICI. By systematically synthesizing these data, we aim to identify the methodological factors driving current outcome heterogeneity and clarify the therapeutic potential of tACS in PD management.

## 2 Results

A comprehensive search of five databases yielded a total of 497 records. Following the removal of 189 duplicates, 308 unique records were retained for title and abstract screening. Although Brittain et al. (2015) ^21^ met most of our inclusion criteria, this study was excluded because the intervention was not a conventional open-loop tACS protocol; instead, stimulation was phase-locked to ongoing tremor and used tremor activity as feedback to determine stimulation timing, making it a closed-loop/phase-synchronised design. Ultimately, ten studies encompassing 184 patients with PD met the inclusion criteria, evaluating the effects of tACS on motor symptoms and neurophysiological motor function (Figure 1). Of these, nine were randomized controlled trials (RCTs)—comprising two parallel-group and seven crossover designs—and one was a non-randomized study. Notably, four of the ten included studies were conducted by the same first author. The pooled cohort consisted of 61 females and 123 males, with an overall mean age of 64.86 years. Due to missing variance data in Shill et al. (2011) ^22^, the overall standard deviation (SD) for age was (imputed with the average SD of 9.4) at 10.33 years (with a lower bound of 9.78 years, assuming zero variance for the missing data). The mean disease duration was 4.93 ± 3.34 years (with average SD of 2.98 imputed for Shill et al. (2011) ^22^). Detailed study characteristics are summarized in Table 1.

**Fig. 1.**
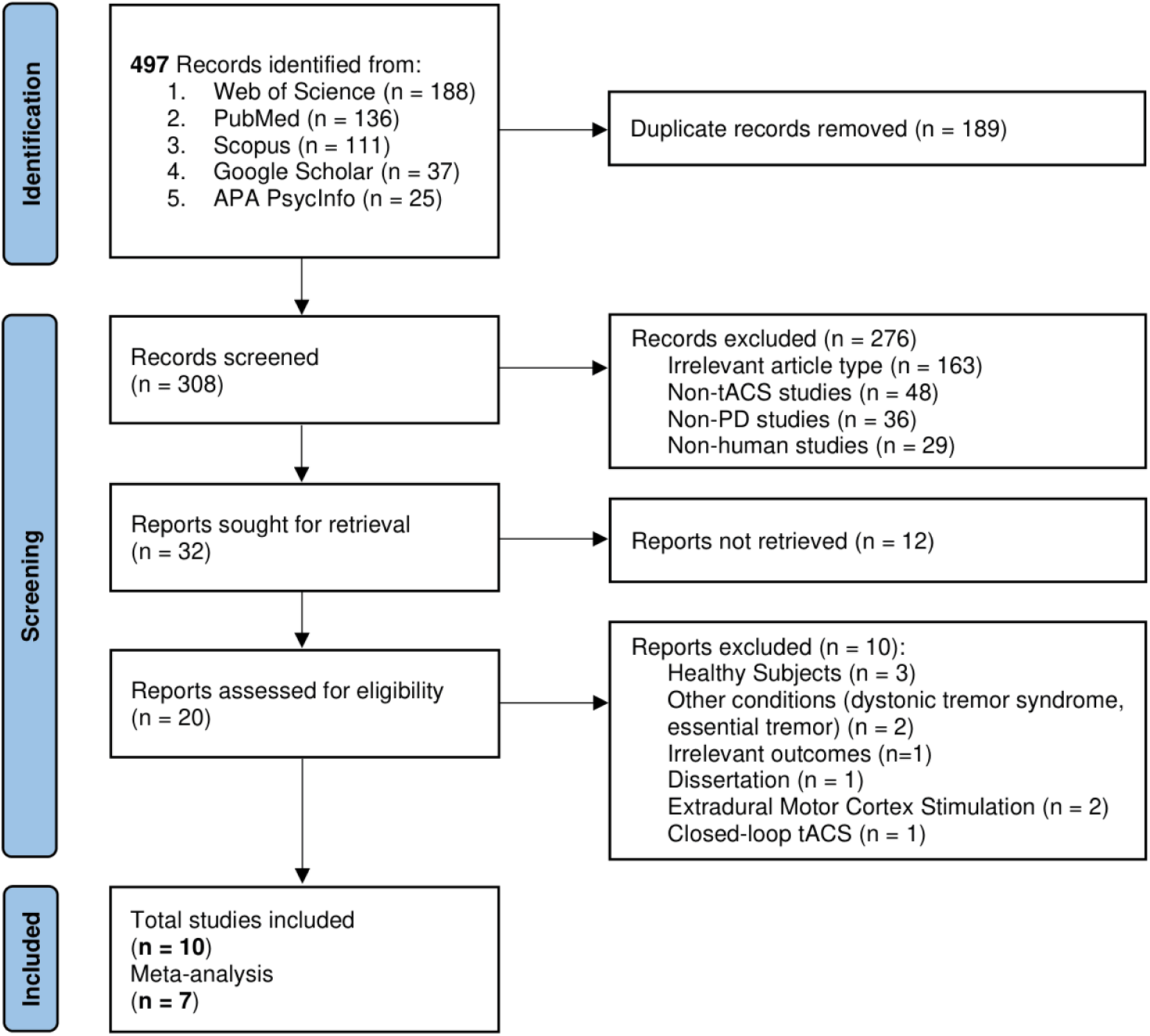
PRISMA flow diagram of the study selection and screening process. The flowchart summarizes the identification of 497 initial records and the systematic exclusion of studies based on predefined eligibility criteria. A total of 10 studies were included for qualitative synthesis, with seven ultimately contributing to the quantitative meta-analysis. n: number of studies.

**Table 1:**
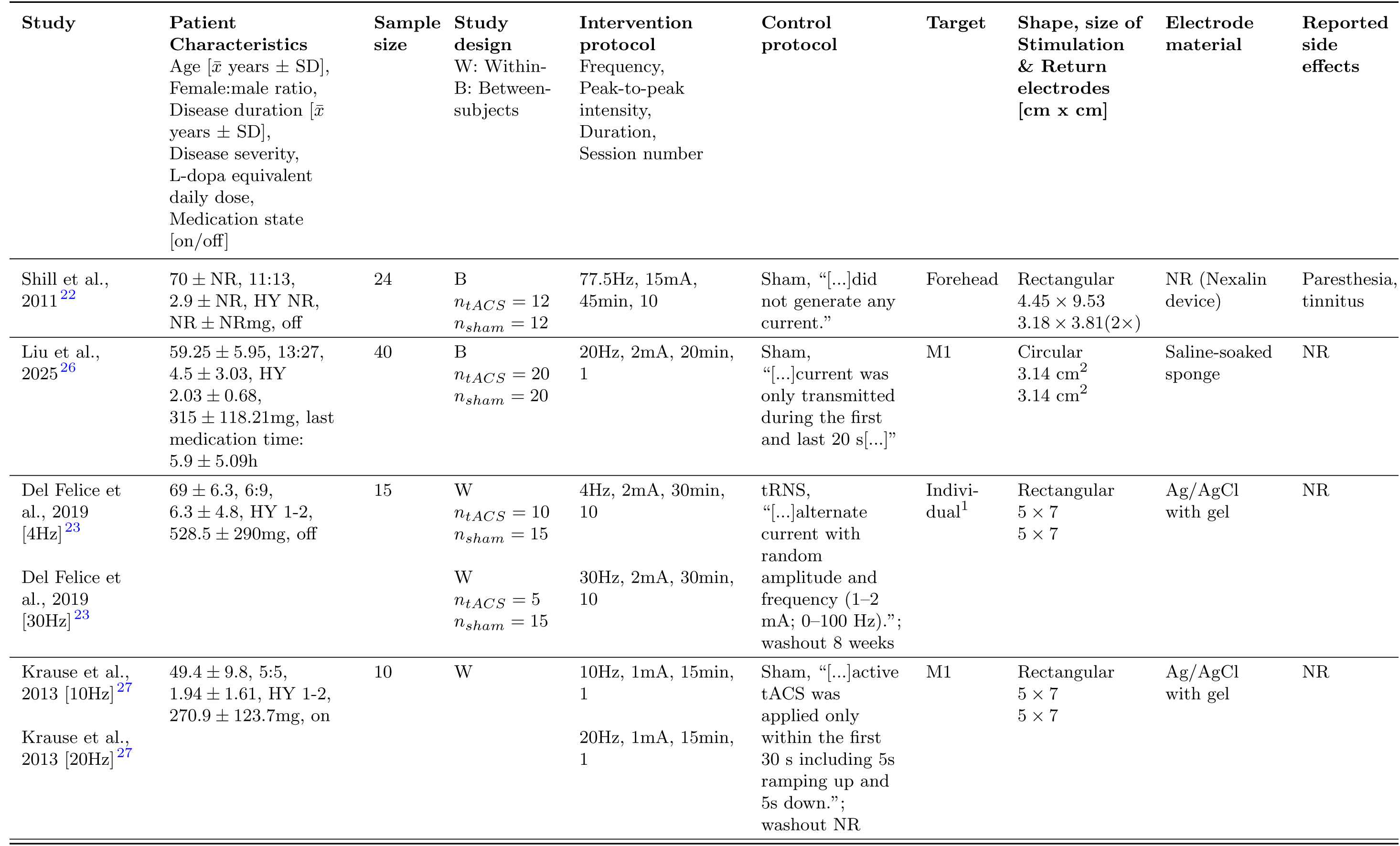
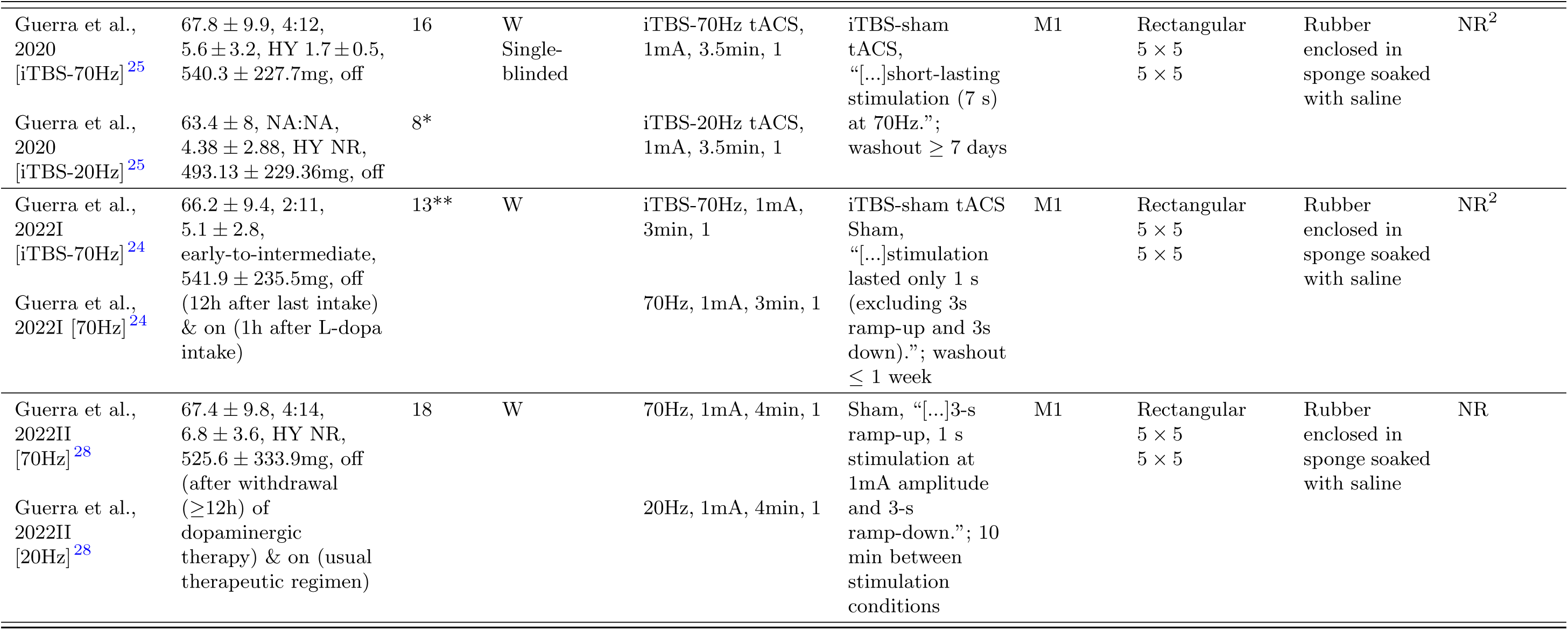
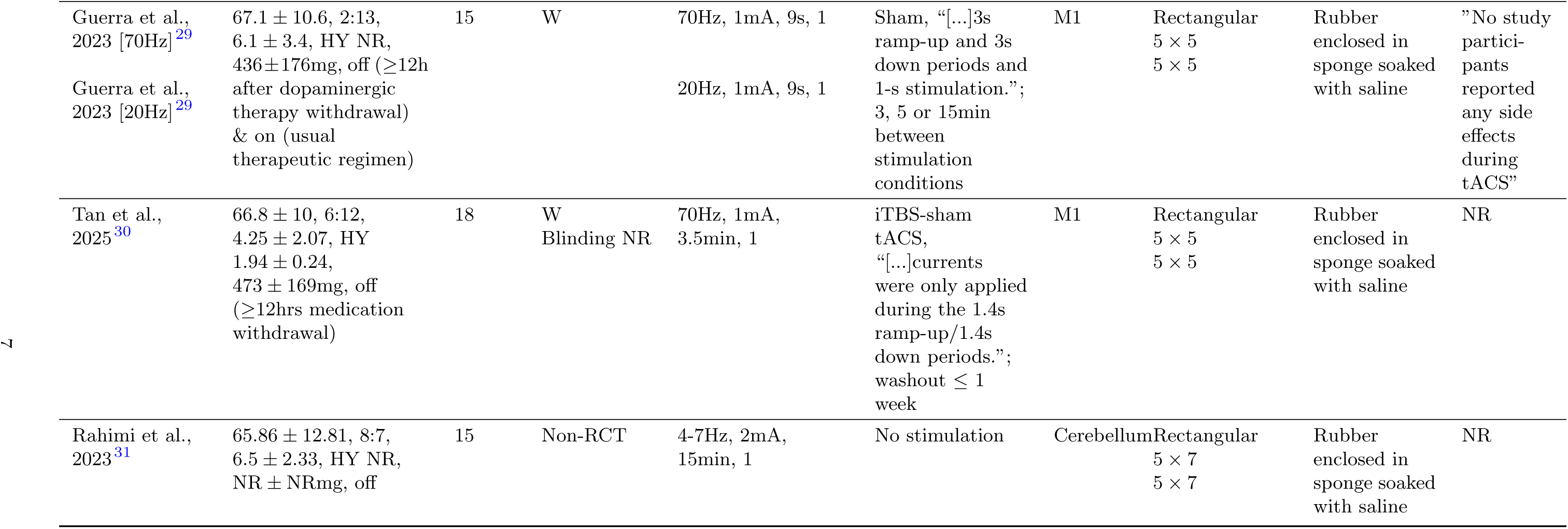
Characteristics of included studies reporting motor outcomes following tACS. *x̄*: mean; SD: Standard Deviation; HY: Hoehn and Yahr stage; NR: Not Reported; tRNS: transcranial Random Noise Stimulation; iTBS: intermittent Theta Burst Stimulation; M1: Primary Motor Cortex; RCT: Randomized Controlled Trial. Unless otherwise specified, all trials utilized a double-blinded RCT design. For extraction transparency, distinct intervention contrasts from the same publication are reported as separate rows. Asterisks (*) denote a subgroup of a larger study cohort, (**) Guerra et al. (2022I) ^24^ states: ”Ten out of 13 patients took part in our previous study on the effect of *γ*-tACS on M1 plasticity in the disease per se (OFF state) ^25^”. ^1^”Electrodes were positioned respectively over the scalp area in which the power spectral difference [between subjects with PD and healthy matched controls] was detected and over the ipsilateral mastoid. ^2^ Guerra et al. (2020) and Guerra et al. (2022I) ^24,25^ did not report specific side effects, however they stated ”[…]tACS did not induce skin or visual sensations in any participant[…]” for the blinding process.

Seven of the ten studies (two parallel-group, four crossover, and one non-randomized) provided sufficient data for inclusion in the meta-analysis. This subset comprised 146 patients with PD (mean age: 65.57 9.6 years; mean disease duration: 4.99 3.37 years). Regarding medication state, five studies assessed outcomes while patients were in the ”OFF” medication state, whereas two studies evaluated patients in both ”ON” and ”OFF” states. Clinical motor symptom outcomes primarily consisted of overall motor severity, assessed via the Unified Parkinson’s Disease Rating Scale (UPDRS; n=3), and specific tremor assessments (n=4). Neurophysiological motor function outcomes predominantly focused on corticospinal excitability, measured using TMS (n=3). Outcome assessments were conducted at various time points: at baseline, during stimulation, immediately and at short-term intervals (15, 20, or 30 minutes) and 3 days post-stimulation ^22^ following single or repeated sessions of theta-, beta-, or gamma-tACS. Extended longitudinal follow-ups were conducted by Shill et al. (2011) ^22^ at 2, 6, 10, and 14 weeks post-stimulation, and by Del Felice et al. (2019) ^23^ at 4 weeks post-stimulation. However, to ensure comparability, the present meta-analyses were restricted to data collected exclusively at baseline (pre-stimulation), during stimulation, and the first timepoint post-stimulation. Tables 2 and 3 provide an overview of assessed outcomes across all 10 studies.

**Table 2.**
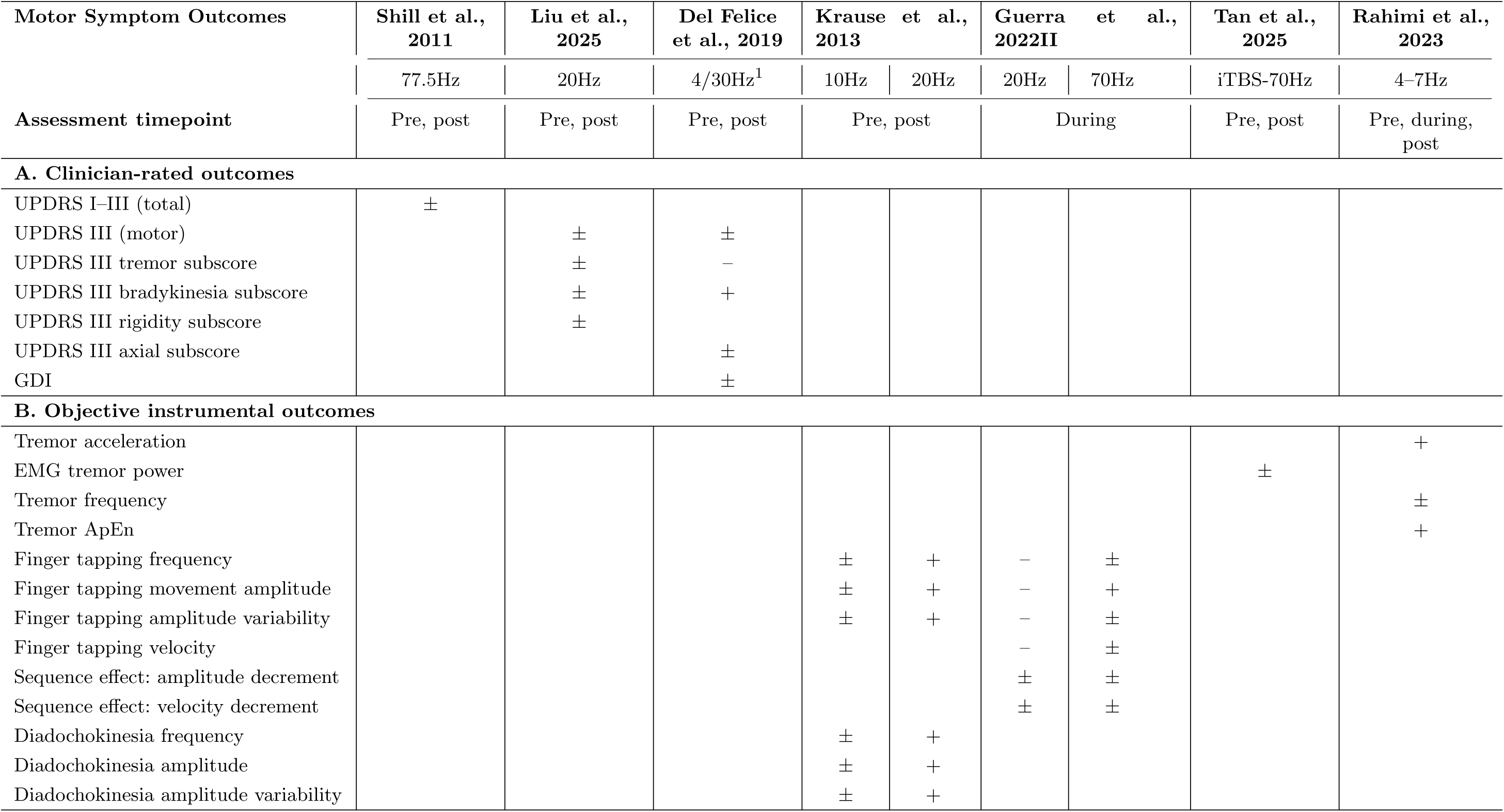
Summary of motor symptom outcomes following tACS, grouped by outcome class. Outcomes are separated into (A) clinician-rated scales and (B) objective instrumental measures to improve interpretability and avoid construct mixing. Symbols indicate within-study direction of reported effect: (+) statistically significant improvement; (–) statistically significant worsening; (*±*) no statistically significant change. ^1^ Del Felice et al. (2019) ^23^ used individualized frequencies (4 and 30 Hz), but frequency-specific outcome effects were not reported separately; effects are therefore shown at the study level. Abbreviations: GDI, Gait Dynamic Index; iTBS, intermittent Theta Burst Stimulation; UPDRS, Unified Parkinson’s Disease Rating Scale; ApEn, Approximate Entropy.

**Table 3.** Summary of neurophysiological motor function outcomes following tACS. The table categorizes observed changes in cortical and corticospinal excitability across varying stimulation frequencies. (+) indicates a statistically significant increase in the respective neurophysiological value; (–) indicates a significant decrease; (*±*) indicates no significant change. Outcomes are primarily derived from Transcranial Magnetic Stimulation (TMS), with additional metrics from Electroencephalography (EEG) and Magnetoencephalography (MEG). Abbreviations: iTBS: intermittent Theta Burst Stimulation; * denotes studies where raw quantitative values were not reported.

| Neurophysiological Motor Function Outcomes | Liu et al., 2025 | Krause et al., 2013 |  | Guerra et al., 2020 |  | Guerra et al., 2022I |  | Guerra et al., 2022II |  | Guerra et al., 2023 |  | Tan et al., 2025 |
| --- | --- | --- | --- | --- | --- | --- | --- | --- | --- | --- | --- | --- |
|  | 20Hz | 10Hz | 20Hz | iTBS-20Hz | iTBS-70Hz | iTBS-70Hz | 70Hz | 20Hz | 70Hz | 20Hz | 70Hz | iTBS-70Hz |
| Assessment timepoint | Pre, post | Pre, post |  | Pre, post |  | Pre, post | During | During |  | During |  | Pre, post |
| TMS Single-Pulse |  |  |  | ± | + | + | + | ± | ± | ±* | ±* | + |
| TMS Short Intracortical Inhibition |  |  |  | ± | + | + | + | − | − | −* | −* | ± |
| EEG power | − |  |  |  |  |  |  |  |  |  |  |  |
| MEG Cortico-Muscular Coupling |  | ± | + |  |  |  |  |  |  |  |  |  |
| TMS Short-latency Afferent Inhibition |  |  |  |  |  |  |  | ± | + |  |  |  |
| TMS Short-Term Potentiation |  |  |  |  |  |  |  |  |  | + |  |  |

### 2.1 Quality of included studies

Overall, most studies demonstrated some concerns regarding the randomization process (Domain 1), primarily due to insufficient reporting of sequence allocation (e.g., failing to specify the exact number of participants assigned to each initial treatment condition) and a low risk of bias for deviations from intended interventions (Domain 2). Among the crossover trials, the two out of 4 raised some concerns regarding potential carryover effects (Domain S). Regarding missing outcome data (Domain 3), four studies were assessed as having a low risk of bias, whereas two randomized trial exhibited some concerns. Bias in the measurement of outcomes (Domain 4) was generally judged to be at low risk. Conversely, the selection of the reported results (Domain 5) introduced some concerns across three studies and a high risk of bias in one study. Consequently, the most significant methodological vulnerabilities across the included literature were observed in the domains of missing outcome data and the selection of reported results. Rahimi et al. (2023) ^31^ was assessed using the 7-domain ROBINS-I tool. It received an overall ”Serious Risk” rating, which was driven entirely by a serious risk of bias due to confounding (Domain 1) and due to deviations from intended interventions (Domain 4), despite scoring low or moderate risk in the remaining domains. While most Del Felice et al, and Liu et al, specifically reported their pre-registrations at ClinicalTrials.gov, all other studies didn’t report it. Detailed risk of bias assessments for each study are presented and summarized in Figure 2.

**Fig. 2.**
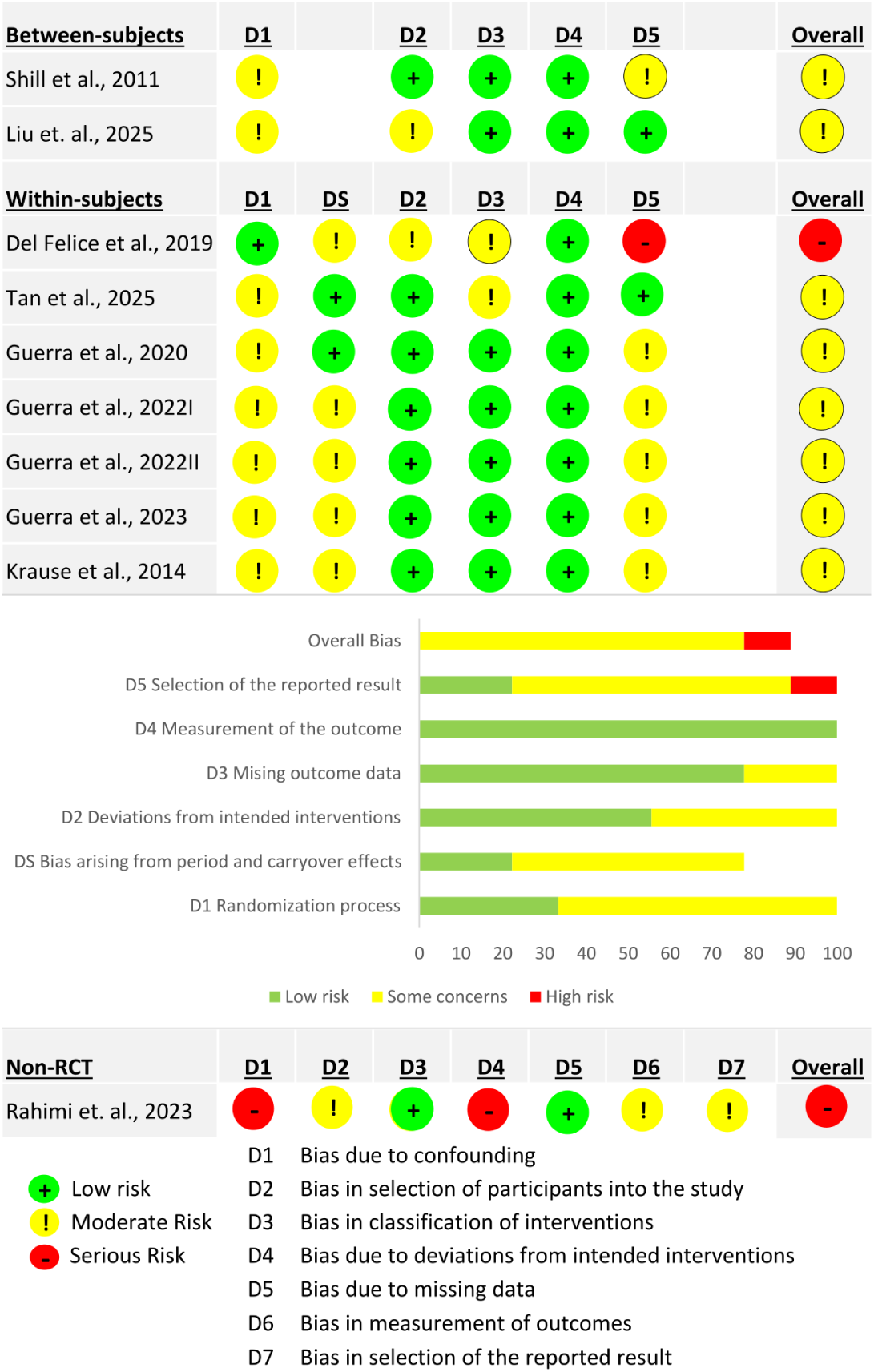
Risk of bias assessment of the included studies. Top and Middle (RCTs): Study-specific evaluations using the Cochrane RoB 2 tool across standard domains (D1–D5) and a supplemental carryover effects domain (DS). The middle bar chart summarizes the overall percentage distribution of risk ratings across all randomized trials. Bottom (Non-RCT): Evaluation of the non-randomized study using the 7-domain ROBINS-I tool. Across all panels, risk levels are color-coded as low (green), moderate/some concerns (yellow), and high/serious risk (red).

### 2.2 Motor Symptom

Figure 3 presents forest plots illustrating the individual and weighted average effect size estimates for the outcome measures of UPDRS score and tremor.

**Fig. 3.**
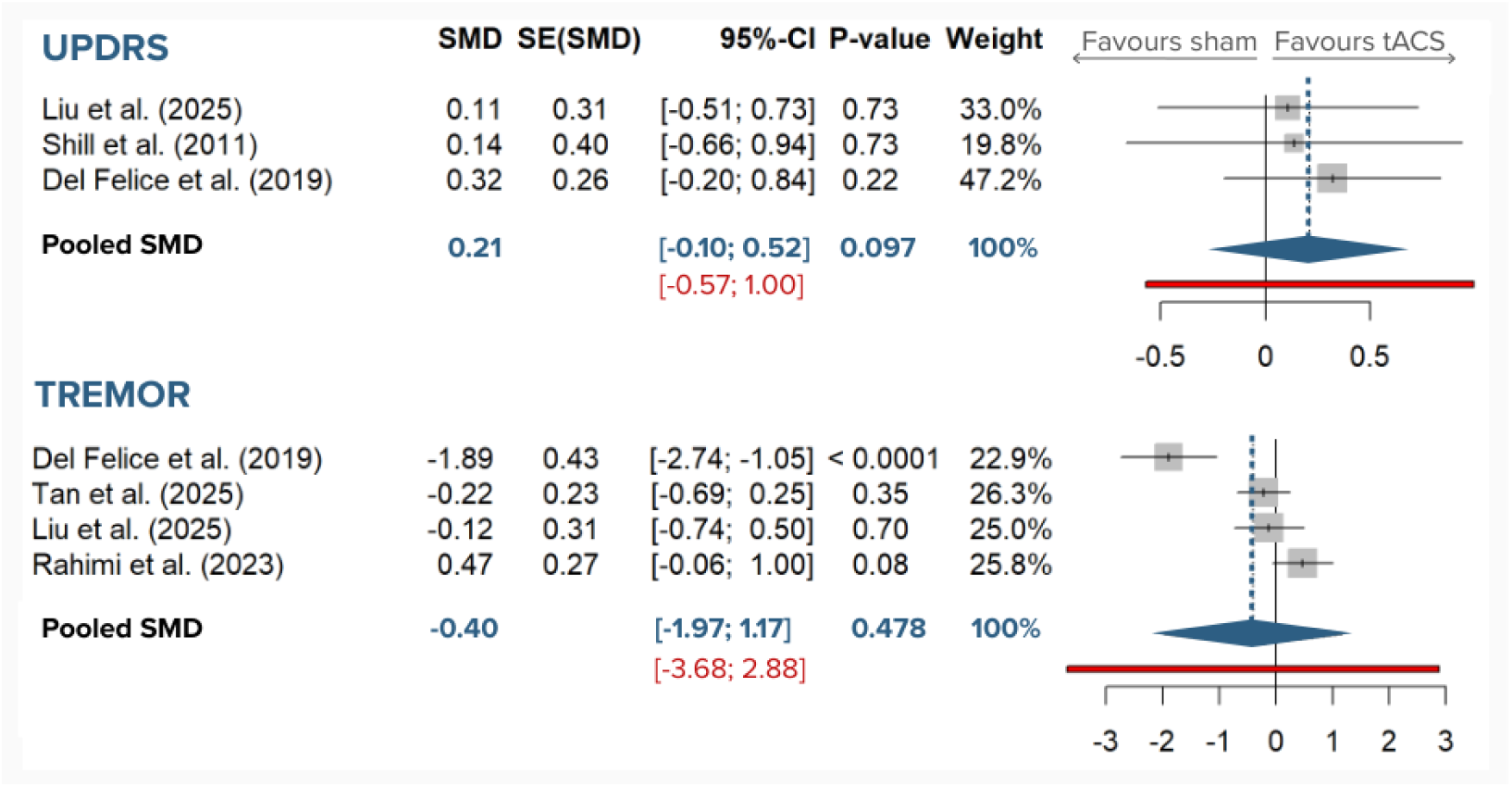
Forest plots of tACS effects on motor symptom outcomes. The plots display the standardized mean differences (SMD) for overall motor severity (UPDRS) and tremor. Individual study effects are represented by squares, with horizontal lines indicating 95% confidence intervals (CI). The pooled effect estimate from the random-effects model is shown as a diamond. The red horizontal bars represent the 95% prediction intervals, illustrating the expected range of effects in future studies. Please note that UPDRS and TREMOR have been inverted such that an increase reflects an improvement.

#### 2.2.1 Unified Parkinson’s Disease Rating Scale

Although eight studies reported baseline UPDRS scores, only three reassessed these measures following the tACS intervention. Among these, Liu et al. (2025) ^26^ and Del Felice et al. (2019) ^23^ reported specific UPDRS Part III (motor examination) subscores, whereas Shill et al. (2011)^22^ reported only the combined sum score for Parts I–III. While Part I assesses non-motor experiences of daily living, the combined Parts I–III score is still predominantly driven by motor aspects. Therefore, to synthesize these varying outcome measures, we pooled the data using the standardized mean difference (SMD). To ensure directional consistency across the analysis—where a positive effect size denotes clinical symptom improvement (i.e., a reduction in UPDRS score)—all calculated effect sizes were multiplied by 1.

The pooled analysis of these three trials comprised 47 active and 47 sham tACS arms. Of the 79 unique patients, 15 were evaluated in a crossover framework, receiving both active and sham tACS. Stimulation parameters varied across studies, targeting the forehead at 77.5 Hz (*k* = 1), the M1 at 20 Hz (*k* = 1), and an individualized montage (F3, FC1, FC5, C3, C4, CP5, Pz) at either 4 Hz or 30 Hz (*k* = 1). The pooled effect size (Hedges’ *g*) was 0.21 (95% CI: 0.10 to 0.52; Figure 3). This indicates a trend toward greater clinical motor improvement following active tACS compared to sham; however, this mean difference did not reach statistical significance (*t* = 2.97, *df* = 2, *p* = 0.097). There was no evidence of statistical heterogeneity across the included studies (*τ* ^2^ = 0; *I*^2^ = 0.0%; Cochran’s *Q* = 0.31, *df* = 2, *p* = 0.856). The pooled analysis yielded an *I*^2^ statistic of 0% and a non-significant Cochrane’s Q test. With only three contributing trials (k = 3), these pooled estimates should be considered exploratory. The absence of detected statistical heterogeneity should not be interpreted as evidence of true homogeneity, as both the *I*^2^ statistic and Cochran’s Q test have very low power to detect heterogeneity when the number of studies is small. Nevertheless, the 95% prediction interval (PI: 0.57 to 1.00) was notably wider than the 95% confidence interval, reflecting uncertainty regarding the true effect in future studies.

#### 2.2.2 Tremor

While Liu et al. (2025) ^26^ and Del Felice et al. (2019) ^23^ reported specific tremor subscores derived from UPDRS III, the remaining two studies utilized physiological metrics to assess tremor. Specifically, Tan et al. (2025) ^30^ measured tremor power via magnetoencephalography (MEG) across three hand muscles; to facilitate comparison, we calculated the grand mean and pooled SDs across these muscles. Rahimi et al. (2023) ^31^ quantified the acceleration of resting tremor, which reflects the overall amplitude and power of the tremor movements. Because all these varied metrics fundamentally assess tremor severity, they were synthesized using the SMD. To maintain directional consistency—where a positive effect size denotes clinical symptom improvement (i.e., a reduction in tremor severity)— all calculated effect sizes were multiplied by 1. Furthermore, data imputation was required for Del Felice et al. (2019) ^23^, who reported the mean UPDRS III tremor subscore without its corresponding SD. Following the retrieval of the overall UPDRS III SD via direct correspondence with the authors, the subscore SD was imputed assuming a proportional variance relationship (i.e., scaling the overall SD by the ratio of the subscore mean to the overall score mean).

The pooled analysis for tremor outcomes incorporated four studies, comprising 68 patients receiving active tACS, 53 receiving sham tACS, and 15 in a no-stimulation control condition. Of those 88 unique participants, 33 received both sham and real tACS in a within-subject design. Stimulation parameters varied across the included studies: targets included M1 at 20 Hz and iTBS-70 Hz (*k* = 2), an individualized montage at either 4 Hz or 30 Hz (*k* = 1), and the cerebellum at 4–7 Hz (*k* = 1). The pooled Hedges’ *g* was 0.40 (95% CI: 1.97 to 1.17; Figure 3) with the overall SMD not being statistically significant (*t* = 0.81, *df* = 3, *p* = 0.48).

Notably, the analysis revealed substantial and statistically significant heterogeneity among the true effect sizes (*τ* ^2^ = 0.83; Cochran’s *Q* = 21.55, *df* = 3, *p <* 0.0001). The *I*^2^ index indicates that approximately 86.1% of the total variance in the observed tACS effects on tremor reflects true between-study variance rather than random sampling error. Consequently, the 95% PI (−3.68 to 2.88) was drastically wider than the 95% CI. A sensitivity analysis demonstrated that excluding Del Felice et al. (2019) ^23^—which exhibited the largest negative effect (*g* = −1.89, weight = 22.9%)— shifted the pooled estimate closer to zero (*g* = 0.04, 95% CI: 0.90 to 0.98; *k* = 3). However, significant heterogeneity persisted during leave-one-out sensitivity analyses (*τ* ^2^ = 0.07 and *I*^2^ = 49.4%), indicating that the observed inconsistency in the literature is robust and not solely driven by a single outlier study.

Given that Rahimi et al. (2023) ^31^ was the only non-randomized pre–post study, we conducted a sensitivity analysis restricted to sham-controlled randomized studies (Liu et al., Del Felice et al., Tan et al.; *k* = 3). In this RCT-only model, the pooled effect remained non-significant (*g* = −0.70, 95% CI: 3.12 to 1.72, *t* = −1.25, *p* = 0.3388), and heterogeneity remained high (*τ* ^2^ = 0.80, *I*^2^ = 84.8%, *Q* = 13.20, *df* = 2, *p* = 0.0014). Thus, exclusion of the non-RCT did not alter the overall inference of uncertain and highly heterogeneous tremor effects.

### 2.3 Neurophysiological Motor Function

Of the ten included studies, seven reported at least one neurophysiological motor function outcome; however, quantitative synthesis was only feasible for four of these studies which assessed inhibitory response using a PP TMS protocol. Notably, three of these investigations were conducted by the same first author with Guerra et al. (2020) ^25^ and Guerra et al. (2022II) ^24^ comprising 10 identical patients. Here, we only included Guerra et al. (2020) ^25^ into the synthesis because of the larger sample size (16 in comparison to 13 in Guerra et al. (2022II) ^24^). Tan et al. (2025) ^30^ employed a highly comparable experimental design. Across these trials, all interventions targeted the M1. The pooled sample comprised 34 patients who received iTBS-70 Hz tACS (*k* = 2) and 18 patients who received 70 Hz tACS (*k* = 1), alongside the same patients assigned to corresponding sham conditions (iTBS-sham or sham). Because the included studies employed standardized TMS protocols to quantify MEP amplitudes, the absolute mean difference (MD) was used to pool the overall effect sizes. To align the directionality of SICI outcomes with clinical improvement, the estimated study-specific effect sizes for SICI were multiplied by 1. In the source publications, SICI was reported as a ratio, calculated by dividing the amplitude of the conditioned MEP (elicited by the PP protocol) by the unconditioned test MEP (elicited by a SP). Physiologically, a decrease in this ratio indicates a smaller conditioned MEP, which reflects stronger intracortical inhibition and enhanced GABA_A_ activation. Therefore, applying this directional correction ensures that a positive pooled effect size correctly represents a therapeutically favorable increase in intracortical inhibition.

The overall pooled MD was 0.00 (95% CI: 0.40 to 0.41; Figure 4). This MD was therefore not statistically significant (*t* = 0.04, *df* = 2, *p* = 0.971), indicating no robust differential effect of active tACS on SICI compared to sham. Effect sizes were completely homogeneous across the included studies (*τ* ^2^ = 0; *I*^2^ = 0.0%; Cochran’s *Q* = 0.83, *df* = 2, *p* = 0.660) acknowledging the limitation of only 2 degrees of freedom. The calculated 95% PI (−0.63 to 0.63) closely mirrored the 95% confidence interval, again highlighting similar bounds of uncertainty for true effects in subsequent studies. To evaluate whether the inclusion of iTBS-primed protocols influenced the pooled estimate, we conducted a sensitivity analysis separating the two iTBS-*γ*-tACS studies ^25,30^ from the standalone *γ*-tACS study ^28^. The pooled MD for iTBS-*γ*-tACS studies alone was 0.09 (95% CI: -0.36 to 0.53; *k* = 2), while the standalone study yielded an MD of 0.20 (95% CI: 0.73 to 0.33). These subgroup estimates did not materially differ from the overall pooled estimate, though the small *k* precludes formal subgroup comparison.

**Fig. 4.**
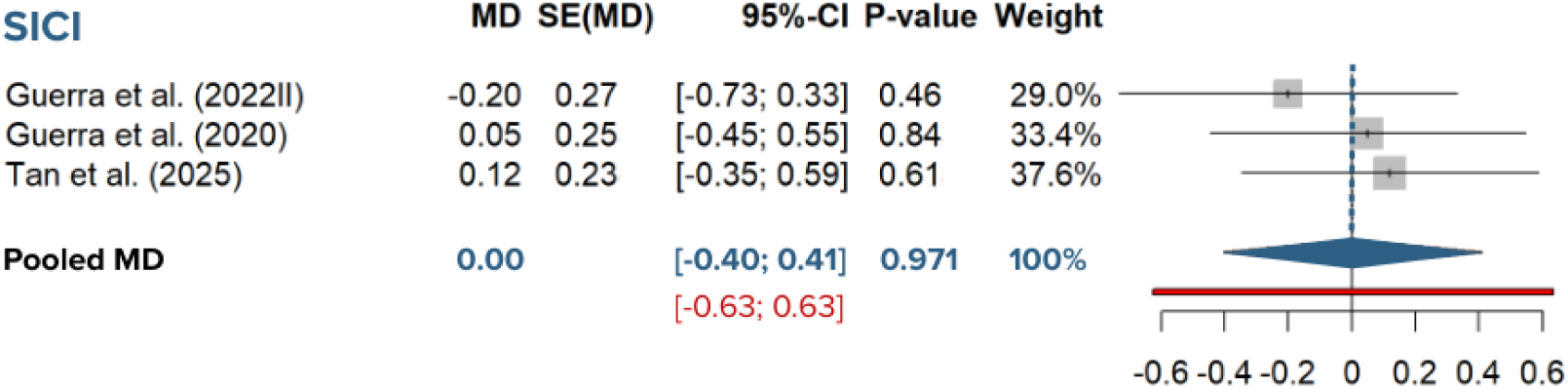
Forest plots of tACS effects on neurophysiological motor function. The plots illustrate the mean differences (MD) short intracortical inhibition (SICI) following transcranial magnetic stimulation. Individual study estimates are indicated by squares with associated 95% confidence intervals (CI). Pooled estimates from the random-effects model (diamonds) are presented alongside 95% prediction intervals (red bars). Positive values indicate an increase in inhibition favoring tACS relative to sham.

Beyond UPDRS, tremor, and SICI, included studies reported numerous additional clinical and instrumental outcomes (Tables 2 and 3). These outcomes are presented as an overview to document the breadth and heterogeneity of endpoint selection across studies. Because constructs, acquisition methods, and reporting formats were highly diverse, these outcomes were not quantitatively pooled and are interpreted descriptively only. A formal GRADE Summary-of-Findings assessment was not performed due to sparse outcome-specific datasets and extensive methodological heterogeneity; however, certainty was judged narratively as low to very low based on risk of bias, inconsistency, and imprecision.

## 3 Discussion

This systematic review and meta-analysis synthesized current evidence on the effects of tACS on motor symptoms and neurophysiological motor function in patients with PD. Across the ten included studies, our meta-analysis of seven trials revealed no statistically significant improvements in overall motor severity (measured via UPDRS), tremor, or SICI.

Although the pooled UPDRS estimate suggested a small favorable trend (*g* = 0.21), this effect was imprecise and accompanied by wide uncertainty intervals. Importantly, effects of this magnitude may be difficult to distinguish from natural intra- and inter-individual symptom variability in PD, and therefore their immediate clinical relevance remains uncertain. At the same time, small average effects should not be dismissed as inherently irrelevant in this field. First, clinically meaningful changes may be symptom-domain specific and therefore diluted in aggregate endpoints such as total UPDRS III. Second, modest pooled effects across heterogeneous cohorts may mask larger benefits in biologically distinct responder subgroups. Finally, given the favorable tolerability profile of tACS, even small but reproducible adjunctive effects could become clinically relevant if confirmed in adequately powered, protocol-specific trials.

For UPDRS, the apparent absence of statistical heterogeneity (*I*^2^ = 0%) should not be over-interpreted. With only three contributing studies, heterogeneity tests are underpowered; therefore, non-detection of heterogeneity is not evidence of true homogeneity. Prediction intervals were consistently broader than confidence intervals and are particularly informative in this setting. While confidence intervals quantify uncertainty around the mean pooled effect, prediction intervals estimate the range of effects that may occur in a new study and therefore better capture between-study variability. The wide prediction intervals observed here indicate substantial uncertainty in both effect magnitude and direction across future PD cohorts, limiting immediate clinical generalizability. At present, the most consistent conclusion is that tACS appears feasible and generally well tolerated, whereas efficacy remains inconclusive. This aligns with the broader literature, which establishes a strong safety profile for tACS devoid of serious adverse effects ^32,33^. Within the current review, the majority of included studies did not report any adverse events, and those that did noted only minor, transient side effects. Furthermore, the implicitly low dropout rates across trials suggest that patient attrition due to treatment-related discomfort is minimal. Nevertheless, while the safety profile of tACS is promising, its clinical and neurophysiological efficacy remains inconclusive based on the current body of evidence.

A primary challenge highlighted by our findings is the measurement of clinical motor outcomes. The UPDRS Part III is the gold standard for assessing motor symptom severity; however, our results raise the question of whether the *overall* UPDRS III score is sensitive enough to capture the nuanced effects of tACS. PD motor symptoms—such as resting tremor, bradykinesia, and rigidity—are mediated by overlapping but distinct pathophysiological network dysfunctions. Because tACS is often tuned to specific frequencies (e.g., beta-tACS for bradykinesia), an aggregated UPDRS III score may dilute domain-specific improvements. For instance, our systematic search found that only a fraction of studies reported subscores (e.g., Liu et al. (2025) ^26^; Del Felice et al. (2019) ^23^). To accurately capture targeted symptomatic changes, it is imperative that future studies not only utilize the UPDRS III assessment pre- and post-tACS but consistently report its specific subscores. Furthermore, to achieve an even deeper and more objective understanding of these clinical effects, future trials should follow the example of Krause et al. (2013) ^27^, Guerra et al. (2022II) ^28^, and Rahimi et al. (2023) ^31^ by integrating kinematic tools, such as wearable movement sensors, to continuously quantify fine motor changes.

Because the precise mechanisms by which tACS modulates motor behavior and motor-related neurophysiology in PD are not fully understood, detailed reporting of neurophysiological data is crucial. In this regard, it is helpful to contrast the trajectory of tACS research with that of DBS. When DBS was first introduced, its clinical effects—such as immediate tremor arrest—were so robust and observable that clinical readouts were sufficient to drive its adoption. It took decades of subsequent research to begin untangling the underlying mechanisms of DBS ^10,34^, a process that is still ongoing today. However, tACS cannot be approached with the same paradigm. The clinical effects of tACS are far more subtle and inconsistent. Therefore, researchers must put a greater effort into incorporating neurophysiological methods (such as EEG, MEG, and TMS, as seen in the studies by Krause et al. (2013) ^27^ and Guerra et al. (2020, 2022I, 2022II, 2023) ^24,25,28,29^) to prove target engagement and elucidate the underlying mechanistic effects.

A key interpretational issue is target plausibility. Much of the field has been motivated by models of subcortical hypersynchronization; however, conventional scalp tACS is biophysically better positioned to modulate cortical rather than deep subcortical activity. Therefore, modest and heterogeneous effects may not only indicate limited efficacy, but also a target-mechanism mismatch in some protocols. In this context, cortical compensatory circuits may represent a more physiologically accessible and potentially more appropriate target class for tACS than attempts to indirectly normalize deep subcortical generators.

The core problem in the current state of tACS research in PD lies in the profound heterogeneity across three main pillars:

1. **Sample characteristics:** Included cohorts exhibited significant heterogeneity in baseline disease severity and duration (range: 0.3 to 10.4 years), as well as pharmacological management, with L-dopa Equivalent Daily Doses ranging from 147.2 mg to 777.4 mg. Notably, while nine of the ten studies assessed patients in a defined ‘OFF’ medication state, reporting standards for the withdrawal period varied; only five studies specified the exact duration of medication withdrawal, while the remaining four omitted this critical temporal detail. Dopaminergic therapy fundamentally alters cortical excitability profiles ^7^ and beta-band synchronization ^3^— one of the oscillatory targets of tACS—and may therefore modulate the responsiveness of cortical circuits to external stimulation. Although formal moderator analysis was not feasible with the current *k*, future meta-analyses with a larger evidence base should examine medication state as a potential effect modifier.
2. **Stimulation protocols:** There is currently no standardized tACS protocol for PD. Our review identified vast differences in stimulation frequencies (ranging from 4 Hz to 77.5 Hz), peak-to-peak intensities (1 to 15 mA), target montages (M1 vs. cerebellum vs. individualized areas), electrode sizes, the number of sessions and duration of each session applied (ranging from 9 seconds to 45 minutes). Interestingly, while the majority of studies utilized a conventional sham protocol— where no current was delivered for most of the stimulation period—Del Felice et al. (2019) ^23^ employed an active control protocol using transcranial random noise stimulation (tRNS) (Table 1). Notably, this study also yielded the largest (positive or negative) SMDs for both UPDRS and tremor outcomes (Fig. 3), suggesting that the choice of the control condition may substantially influence the magnitude of the observed therapeutic effects.
3. **Outcomes assessed:** As noted, the endpoints varied so widely that only a fraction of the identified studies’ outcomes could be pooled for meta-analysis (Tables 2 and 3). Although objective motor outcomes were not meta-analyzed due to methodological non-comparability across studies, they remain highly relevant. Compared with global clinical scales, kinematic and instrumental endpoints may be more sensitive to subtle, domain-specific motor effects of tACS and may better capture early target-related changes. Their current heterogeneity therefore should not be interpreted as lack of importance, but as a signal that future trials need harmonized objective endpoint frameworks to enable clinically meaningful cross-study synthesis. An additional limitation is temporal heterogeneity of outcome assessment. Included studies evaluated effects during stimulation, immediately after stimulation, minutes later, or several days later, which may reflect different physiological mechanisms (online entrainment vs offline after-effects).

Beyond these protocol-level parameters, the biophysical dose delivered to cortical targets likely varied substantially across studies. Computational modeling estimates suggest that conventional tACS at 1–2 mA peak-to-peak generates cortical electric fields on the order of 0.2–0.5 V/m, well below the approximately 1 V/m threshold typically associated with reliable neural entrainment in animal models ^35^. The 15 mA protocol used by Shill et al. (2011) ^22^ would produce considerably stronger fields, though safety and tolerability at such intensities remain debated. Recent primate work by Tran et al. (2026) ^36^ demonstrated that tACS effects are highly dose-dependent: stronger electric fields entrain motor-cortex neurons to the stimulation rhythm, whereas weaker fields may paradoxically disrupt pathological synchrony by shifting spike timing and preferred phase. This dose-response complexity further complicates cross-study comparison and underscores the need for computational dosimetry reporting in future tACS trials.

Contributing to this heterogeneity is the highly individualized response of each subject to tACS. The physiological rationale for individualized tACS stems from the pathophysiology of PD, which is characterized by hypersynchronized cortical oscillations—particularly within the beta band (13–30 Hz)—that drive motor symptoms ^2,3^. Inter-individual variability in baseline neurophysiology, cortical anatomy, and symptom dominance means that a ”one-size-fits-all” stimulation protocol is likely ineffective. Rather than applying fixed frequencies and intensities, identifying a patient’s specific pathological oscillatory peak (e.g., via resting-state EEG or MEG) and accounting for their distinct cortical anatomy (e.g., disease-related atrophy) allows for precision-targeted delivery. However, because the mechanistic effect of tACS is highly frequency-dependent ^36^, this precision targeting carries a unique risk: when the stimulation frequency exactly matches the brain’s endogenous oscillation, the risk of reinforcing (rather than suppressing) pathological synchronization increases—a fundamental limitation of conventional open-loop entrainment. While Del Felice et al. (2019) ^23^ pioneered an individualized protocol that yielded clinical improvements, the lack of frequency-specific outcome reporting precludes definitive conclusions regarding how personalized parameters influenced these gains.

To effectively desynchronize pathological activity and overcome the limitations of open-loop entrainment, the field must transition toward closed-loop tACS paradigms. These dynamically adaptive protocols offer a promising avenue for personalized therapy by modulating stimulation parameters in real-time based on the brain’s ongoing oscillatory state. While conventional tACS modulates superficial cortical activity, PD pathophysiology often requires targeting deep subcortical networks. To bridge the gap between superficial tACS and invasive DBS, emerging non-invasive techniques like transcranial temporal interference stimulation employ intersecting alternating currents to stimulate deep brain structures ^37^. Integrating such spatially precise, deep-focal methodologies with closed-loop EEG represents the next critical evolution in non-invasive therapeutics for oscillopathies like PD.

The interpretation of our findings must also be contextualized within the methodological quality of the primary literature. As detailed in Section 2.1, despite robust outcome measurements, significant vulnerabilities remain regarding randomization, selective reporting, and blinding integrity—a pervasive challenge in NIBS research. Furthermore, the prevalence of crossover designs raises concerns regarding potential carryover effects. Although the majority of our seven included crossover trials reported washout periods (Krause et al. (2013) ^27^ being a notable exception), the risk of carry-over effects cannot be entirely dismissed due to a lack of standardized ”session doses”. Research on the after-effects of tACS remains sparse, particularly in the beta and gamma ranges, and evidence suggests these effects are highly state-dependent ^38,39^. Consequently, the development of protocols that account for the patient’s immediate physiological state and previous stimulation history is essential. Other specific methodological caveats exist within the pooled studies. While Del Felice et al. (2019) ^23^ reported the largest effect size for UPDRS-III, this study combined tACS with concurrent physical exercise. Because physical exercise is independently known to robustly alleviate motor symptoms in PD ^40^, the isolated efficacy of tACS in this trial remains ambiguous, likely inflating the slight positive trend observed in our UPDRS scores. Additionally, a limitation specific to our tremor synthesis is the pooling of outcome measures that represent partially distinct physiological and clinical constructs (clinical subscores vs. continuous MEG-derived spectral power). A sensitivity analysis restricted to the two studies reporting UPDRS-derived tremor subscores yielded a pooled SMD of -0.98 (95% CI: -12 to 10); however, with only two studies, this estimate should be interpreted with extreme caution. Importantly, the SICI meta-analysis pooled standalone *γ*-tACS with iTBS-*γ*-tACS combination protocols. While all studies utilized comparable TMS assessment paradigms and targeted M1 at gamma frequencies, iTBS itself induces LTP-like plasticity changes that may interact with subsequent tACS effects in ways that are not additive. Although our sensitivity analysis did not reveal meaningful differences between these subgroups, the small number of studies limits the inferential value of this comparison, and future work should assess these protocols separately.

Beyond the primary literature, this meta-analysis is subject to several inherent limitations. First, given the limited number of studies entering each pooled analysis (*k* = 3–4), the present meta-analyses are best interpreted as exploratory syntheses rather than confirmatory tests of tACS efficacy. Post-hoc power calculations indicate that with the observed pooled sample sizes (*N* = 80–146), only large effect sizes (*d* ≥ 0.6–0.8) could be reliably detected at conventional thresholds (*α* = 0.05, 1 − *β* = 0.80), meaning clinically meaningful but moderate effects may have gone undetected. Second, incomplete data reporting in the original manuscripts necessitated the conservative imputation of variance data for certain trials (e.g., Shill et al. (2011) ^22^; Del Felice et al. (2019) ^23^), which inherently introduces a degree of uncertainty into the pooled effect sizes. Finally, to ensure methodological comparability, we strictly restricted our quantitative synthesis to the first reported post-stimulation (or during-stimulation) timepoints; therefore, the long-term therapeutic durability and after-effects of tACS remain largely uncaptured in this analysis.

A recent meta-analysis by Ye et al. (2025) ^20^ reported large positive effect sizes for MEP (SMD = 2.65) and SICI (SMD = 1.88) following tACS in PD. In contrast, the present investigation—utilizing a broader search up to August 2025—found no significant differences between active tACS and sham for any clinical or neurophysiological outcomes. This divergence likely stems from several distinct methodological choices prioritized here to isolate true therapeutic efficacy and ensure physiological accuracy.

The analytical framework herein strictly compares active tACS against sham or no-stimulation exclusively within idiopathic PD cohorts. This design isolates the specific therapeutic effect of the intervention. Comparing PD patients to healthy controls (HCs), or including non-idiopathic pathologies (e.g., dementia with Lewy bodies), primarily addresses disease-related physiological differences rather than evaluating standard clinical efficacy. Furthermore, restricting the control condition to sham stimulation prevents the mathematical distortion of effect sizes. Because HCs exhibit normal MEP ranges that may respond differently to tACS than PD patients, using HCs as a comparative baseline severely complicates the interpretation and directionality of neurophysiological outcomes.

Although Guerra et al. (2020) ^25^, Guerra et al. (2022II) ^28^, and Tan et al. (2025) ^30^ all utilized SP TMS to quantify MEP amplitudes under *γ*-tACS, they measured fundamentally different phenomena. Consequently, reported SP measurements were excluded from pooled synthesis to strictly separate evaluations of *baseline corticospinal excitability* (assessed via fixed-intensity SP MEPs without a preceding induction protocol, as in Guerra et al. (2020) ^25^) from assessments of *LTP-like synaptic plasticity* (post-iTBS MEP facilitation, as in Guerra et al. (2022II)^28^ and Tan et al. (2025) ^30^). Treating moment-to-moment excitability and plasticity capacity as equivalent in a pooled synthesis introduces a significant methodological confound that the current review avoids.

Finally, this analysis strictly accounts for sample overlap to satisfy the core statistical assumption of independence. For instance, data from Guerra et al. (2020) ^25^ and Guerra et al. (2022II) ^24^ involve overlapping participant cohorts; appropriately handling these populations prevents the double-counting of subjects, which can otherwise artificially narrow confidence intervals and bias effect size estimates.

By implementing these rigorous methodological controls—transparent data extraction, separation of distinct physiological metrics, and strict adherence to PRISMA guidelines—the present findings provide a conservative, clinically grounded estimate of the current utility of tACS in the management of PD.

## 4 Conlusion

Taken together, tACS showed a beneficial safety profile. However, there is a lack of consistent improvements in motor symptoms or functions in PD due to small study numbers and large outcome heterogeneity of included studies. The broad prediction intervals limit confidence in a stable treatment effect. Accordingly, these pooled estimates should be interpreted as exploratory and hypothesis-generating rather than confirmatory evidence of efficacy. Accordingly, the present data do not demonstrate protocol-level efficacy or inefficacy; rather, they indicate that evidence is currently insufficient to establish efficacy for any specific tACS protocol in PD. Given substantial heterogeneity in stimulation parameters, patient characteristics, outcome definitions, and assessment timing, definitive conclusions about clinical benefit are premature. To overcome the limitations of the current literature, future tACS trials in PD must adopt a more standardized and transparent approach. Clinically, trials should uniformly assess and report UPDRS III subscores pre- and post-stimulation to allow for the meta-analysis of symptom-specific effects. Mechanistically, moving away from “one-size-fits-all” fixed-frequency protocols toward personalized, closed-loop tACS—guided by concurrent EEG or MEG to match the patient’s individual peak oscillatory frequencies—may prove vital in overcoming the high inter-individual variability and may add to a clearer picture. Additionally, rigorous sham-control procedures with explicit reporting of blinding success are necessary to ensure the validity of both clinical and neurophysiological outcomes.

## 5 Methods

This systematic review and meta-analysis was conducted in accordance with the established Preferred Reporting Items for Systematic Reviews and Meta-Analyses (PRISMA) reporting guidelines (see Supplementaries PRISMA 2020 checklist.docx) and adhered to the preregistered protocol on PROSPERO (Registration Number: CRD420251110394).

### 5.1 Search Strategy

To ensure high sensitivity and comprehensive, cross-disciplinary coverage of the rapidly evolving literature, we selected five complementary electronic databases. **PubMed/MEDLINE** and **Web of Science** provided the core of peer-reviewed biomedical and clinical trial evidence. To account for the neuropsychological and cognitive dimensions of PD, **APA PsycInfo** was included as a specialized behavioral resource. **Scopus** was utilized to broaden the interdisciplinary scope, capturing relevant bioengineering and technical reports often underrepresented in strictly clinical databases. Finally, **Google Scholar** served as a high-sensitivity tool to identify grey literature and recently published reports not yet indexed in traditional repositories.

The search strategy was peer-reviewed by a librarian, and suggested revisions to keyword syntax and Boolean structure were incorporated before execution. We conducted a systematic search from inception to August 31, 2025, utilizing the following tailored search strings:

- **Web of Science & PubMed/MEDLINE:** `(((transcran* AND stimula* AND alternat*) OR tacs) AND parkinson*)`
- **Scopus (utilizing proximity operators):** `(((transcran* W/3 stimula*) W/3 alternat*) OR tacs) AND parkinson*`
- **APA PsycInfo (Filter: Human):** `(((transcran* N3 stimula*) N3 alternat*) OR tacs) AND parkinson*`
- **Google Scholar (incorporating exclusion filters):** `("transcranial alternating current" OR tACS AND "Parkinson") -animal -rat -rats -mouse -mice -"in vitro" -review -editorial -letter -protocol -abstract -poster -dissertation -healthy -schizophrenia`

Additionally, we performed a manual backward citation search of the reference lists from all included studies and relevant prior reviews to identify further eligible records. No automation tools were employed during the search or selection process.

### 5.2 Selection Criteria and Process

The study selection process was conducted in two sequential stages using the Rayyan platform. No automation tools were employed during the process. First, two authors (TTM and TG) independently screened the titles and abstracts of all retrieved records against the predefined **PICOTS** eligibility criteria.

- **Population (P):** Adults diagnosed with idiopathic PD of any age, sex, disease duration, or Hoehn–Yahr stage. Studies were included regardless of baseline medication status (on- or off-dopaminergic therapy) or common comorbidities (such as depression). We excluded atypical or secondary parkinsonism (e.g., drug-induced parkinsonism) and studies involving active DBS unless the device was inactive during tACS and PD-specific results were separable.
- **Intervention (I):** Non-invasive tACS delivered via scalp electrodes, with no restrictions on frequency, intensity, waveform, or session duration. Protocols involving tACS delivered alone or concomitantly with other therapies (e.g., physiotherapy) were included, provided the tACS effect could be analytically isolated. We excluded invasive alternating-current approaches and protocols lacking an alternating-current component.
- **Comparator (C):** RCTs utilizing parallel-group or crossover designs comparing active Tacs against sham (placebo), active-sham, or usual care. Non-RCTs with pre–post designs were also eligible. Given the limited number of sham-controlled randomized trials in this field, we included both randomized controlled trials and non-randomized pre–post studies to comprehensively characterize the available evidence. We then performed a sensitivity analysis only including sham-controlled studies.
- **Outcomes (O):** Primary outcomes were changes in clinician-rated motor severity (e.g., MDS-UPDRS-III), functional motor abilities (objective gait, balance, or dexterity tests), and neurophysiological measures (indices derived from EEG, MEG, or TMS). Secondary outcomes included patient-reported motor function (MDS-UPDRS-II, PDQ-39), motor fluctuations (“on”/“off” time), and safety/adverse event reporting.
- **Timing (T):** Studies were included if they assessed outcomes during the tACS intervention or immediately following its completion. While longitudinal follow-up assessments were documented, they were not a prerequisite for inclusion.
- **Setting (S):** Clinical and research environments including both inpatient and outpatient settings.

Studies were excluded if they involved non-PD populations or PD patients with severe uncommon physical or psychiatric comorbidities. Additionally, non-English and non-German publications, animal models, in vitro research, and non-empirical article types (e.g., reviews, meta-analyses, abstracts, study protocols) were excluded. To maintain a mechanistically homogeneous sample, studies utilizing closed-loop tACS paradigms were also omitted. Rayyan was employed to manage the records identified, as well as for deduplication. Studies meeting all inclusion criteria but lacking sufficient quantitative data for effect size calculation were retained strictly for qualitative synthesis.

Second, the full texts of potentially relevant articles were retrieved and independently assessed for inclusion by the same two reviewers. In cases of disagreement at either stage, a third author was consulted to reach a consensus. The reasons for excluding full-text articles were documented and are summarized in the PRISMA flow diagram (Fig. 1).

### 5.3 Data Selection and Extraction

To reduce selective extraction bias when multiple eligible data points were available within a study, we applied prespecified hierarchical rules. First, for each outcome domain, we prioritized the endpoint designated as primary by study authors; if no primary endpoint was specified, we prioritized the most clinically interpretable and comparable endpoint across studies. Second, for timing, we used the first immediate post-intervention assessment for the primary synthesis; later follow-ups were retained for sensitivity analyses when feasible. Third, when multiple active stimulation arms shared one sham/control arm, we avoided unit-of-analysis errors by combining active arms using Cochrane-recommended formulas (or selecting the prespecified primary active condition when combination was not feasible). Fourth, in crossover studies, paired active-versus-sham contrasts were preferentially extracted over unpaired approximations.

Data extraction was systematically and independently performed by two reviewers (TTM and TG) for each included study. Any discrepancies were resolved through consultation with a third author. The following variables were captured: (a) study metadata (authors, publication year, study design, sample size, and dropout rates); (b) patient demographics (age, sex ratio) and clinical characteristics (disease severity, medication status, disease duration); (c) detailed tACS parameters (target scalp region, frequency, peak-to-peak intensity, duration, number of sessions, and washout periods); (d) specific outcome measures and instruments utilized to assess motor symptoms and function; (e) quantitative outcome data, including means and SDs for both active and sham conditions. When means and SDs were not directly available, we applied the following conversion methods. In cases where publications reported only relative changes (e.g., percentage of baseline) along with their respective measures of dispersion (SD or SEM), and absolute baseline values were available, we back-calculated the absolute post-intervention values to ensure consistency across the meta-analysis. If studies reported SEM, we converted to SD using *SD* = *SEM n*. WebPlotDigitizer was used to extract data from figures. When means and SDs were not reported (e.g., results provided only as medians/IQRs, *p*-values, or confidence intervals) or essential quantitative data were missing or incompletely reported, we contacted corresponding authors to obtain the required summary statistics using a standardized three-step protocol: an initial email inquiry, followed by a second reminder at two weeks, and a final reminder two weeks thereafter. All successfully retrieved data were subsequently integrated into the analyses. Finally, we extracted (f) qualitative descriptors of the effect direction, categorized as improvement [+], no significant change [*±*], or worsening [–].

### 5.4 Meta-Analysis

While Cochrane guidelines permit meta-analyses with as few as two or three studies, we explicitly frame our quantitative syntheses as *exploratory* rather than confirmatory due to the limited number of available trials. The primary utility of pooling these data is to summarize the current trajectory of the field and contrast it with previous, highly inflated estimates, rather than to establish definitive clinical efficacy.

All meta-analytic computations were conducted using the meta (and metagen) package in R software (version 4.3.3). For continuous clinical outcomes measured across varying scales (e.g., UPDRS, tremor severity), SMDs were calculated using Hedges’ *g*, accompanied by standard errors and 95% CIs. Where available, pre- and post-tACS change scores were utilized to estimate effect sizes. When outcomes were measured only during/after stimulation (without baseline-adjusted change reporting), effect sizes were calculated from (post-)intervention values comparing active tACS versus sham. When both pre- and post-tACS values were available, we calculated change scores within each arm (Δ = post pre) and computed the between-group effect size using these arm-level change scores (active vs sham). For non-randomized pre–post studies, we calculated standardized mean change (within-group pre-to-post). To ensure the comparability of effect size estimates derived from disparate experimental designs, we mathematically accounted for study design (between-vs. within-subjects) during the calculation of Hedges’ *g* ^41^. For crossover trials, within-subject effect sizes require an estimate of the pre-post correlation (r) to correctly compute the standard error. We estimated this correlation using the reported standard deviations and the standard deviations of the change scores ^42^. Sensitivity analyses were conducted at two plausible additional correlations (r = 0.3 and r = 0.7) to evaluate the robustness of pooled estimates to this assumption (Supplementary Table S2). To maximize the utility of the limited available data, diverse measures of tremor severity—ranging from ordinal clinical ratings (UPDRS tremor subscores) to kinematic (accelerometry) and physiological (MEG tremor power) metrics—were pooled using the SMD. Although these varied metrics all fundamentally index tremor severity, they capture partially distinct constructs (subjective clinical severity vs. spectral power vs. movement amplitude). We therefore synthesized them using the SMD to place all measures on a common scale. Readers should note, however, that this construct heterogeneity may contribute to observed between-study variance beyond what is attributable to differences in stimulation protocols or patient characteristics alone. Conversely, for neurophysiological outcomes assessed via uniform methodologies across studies, MDs were calculated ^41^.

Individual effect sizes (Hedges’ *g* or MD) were pooled across studies using an inverse-variance weighting method and analyzed via a random-effects model, which inherently accounts for between-study variance. To avoid the inflation of effect sizes due to redundant data (double counting), we screened for overlapping patient cohorts across studies from the same research groups. When overlapping populations were identified—specifically between Guerra et al. (2020) ^25^ and Guerra et al. (2022II)^24^ where 10 out of 13 participants were identical—we prioritized the study with the larger sample size for the primary meta-analysis. The precision of the pooled estimates was evaluated using 95% CIs, while 95% PIs were calculated to assess the expected range of true effects in future study settings ^43^. Statistical heterogeneity among the true effect sizes was evaluated using Cochran’s *Q* test and Higgins’s *I*^2^ statistic ^44^.

Although meta-regression analyses were planned a priori to explore potential moderators (age, disease severity, duration, and tACS parameters), these could not be reliably performed outcomes with high heterogeneity due to the limited number of studies (k *<* 10) available for each pooled analysis. Sensitivity analyses were conducted utilizing a leave-one-out approach to determine the influence of individual studies on the pooled effect.

### 5.5 Study Quality and Risk of Bias Assessment

For non-randomized studies, the ROBINS-I tool was utilized to evaluate seven domains of bias: (1) bias due to confounding, (2) bias in selection of participants into the study, (3) bias in classification of interventions, (4) bias due to deviations from intended interventions, (5) bias due to missing data, (6) bias in measurement of outcomes, and (7) bias in selection of the reported result. These domains were categorized as “low”, “moderate”, “serious”, or “critical” risk of bias.

To assess the risk of bias due to missing results (reporting bias), we cross-referenced the outcomes reported in the published manuscripts against their respective trial registries and published protocols, where available. Due to the limited number of studies included in each quantitative synthesis (*n <* 10), formal statistical assessment of publication bias via symmetry testing (e.g., Egger’s test) and funnel plots was not performed, as these methods lack sufficient power to distinguish chance from real asymmetry in small samples ^45^. Instead, we qualitatively assessed the risk by evaluating the comprehensiveness of the search strategy—including grey literature via Google Scholar—and identifying any discrepancies between predefined and reported outcomes. Any discrepancies between the two independent reviewers were resolved through discussion and consensus. The entire assessment process adhered rigorously to the Methodological Expectations for Cochrane Intervention Reviews (MECIR) standards.

### 5.6 Data availability

All data generated or analyzed during this study are included in this published article.

## Data Availability

All data produced in the present work are contained in the manuscript.

## Acknowledgements

We would like to thank Oliver Schoenbeck for his valuable contributions to constructing the database search strings, and Valentin Emslander for his statistical and methodological advice in conducting the meta-analyses.

## Author contributions

T.T.M.: Conceptualization, synthesis, statistical analysis, writing - original draft. T.G.: Synthesis, writing - review & editing. K.W.: Conceptualization, writing - review & editing. M.R.: Conceptualization, writing - review & editing. C.S.H.: Conceptualization, writing - review & editing, supervision. All authors read and approved the final manuscript.

## Additional information

**Supplementary information** The online version contains supplementary material available at XXX.

**Correspondence** and requests for materials should be addressed to Thuy Tien Mai.

