## Supplementary_Tables_S1-S2 for "Transcranial alternating current stimulation for Parkinson’s disease: a systematic review and meta-analysis of motor outcomes"

**Mean and Standard Deviations (SDs)**

| **Study** | **Mean Pre-Stim** | | **SD Pre-Stim** | | **Mean Post-Stim** | | **SD Post-Stim** | |
| --- | --- | --- | --- | --- | --- | --- | --- | --- |
|  | Real | Sham | Real | Sham | Real | Sham | Real | Sham |
| Shill et al. (2011) | 27.42 | 29.71 | 4.50 | 3.64 | 19.95 | 22.81 | 3.98 | 3.98 |
| Liu et al. (2025) | 23.80 | 22.05 | 12.86 | 11.56 | 21.17 | 20.71 | 12.11 | 11.06 |
| Del Felice et al. (2019) | 33.29 | 33.18 | 9.27 | 11.67 | 27.36 | 30.54 | 9.70 | 8.30 |
| Liu et al. (2025) | 1.65 | 1.90 | 1.66 | 2.27 | 1.69 | 1.71 | 1.57 | 1.95 |
| Del Felice et al. (2019) | 0.47 | 0.58 | 0.13 | 0.20 | 0.36 | 0.21 | 0.13 | 0.06 |
| Tan et al. (2025) | -0.35 | 0.14 | 1.38 | 2.13 | -0.21 | -0.18 | 1.60 | 2.72 |
| Rahimi et al. (2023) | 0.03 |  | 0.05 |  | 0.01 |  | 0.01 |  |
| Guerra et al. (2020) | 0.61 | 0.57 | 0.22 | 0.20 | 0.54 | 0.55 | 0.23 | 0.21 |
| Tan et al. (2025) | 0.87 | 0.82 | 0.55 | 0.53 | 0.77 | 0.84 | 0.47 | 0.50 |
|  | **Mean During-Stim** | | **SD During-Stim** | |  |  |  |  |
| Guerra et al. (2022II) | 0.79 | 0.59 | 0.26 | 0.21 |  |  |  |  |

**Table S1** Raw mean and standard deviation (SD) values extracted from the included studies for real and sham tACS conditions, reported for pre-stimulation, post-stimulation, and where available during-stimulation measurements. UPDRS, Tremor, SICI.

**Correlation Sensitivity Analyses**

| **r** | **(S)MD** | **95% CI** | **P-value** |
| --- | --- | --- | --- |
| 0.3 | 0.19 | [-0.04; 0.42] | 0.073 |
| 0.7 | 0.25 | [-0.19; 0.70] | 0.135 |
| 0.3 | -0.33 | [-1.69; 1.04] | 0.505 |
| 0.7 | -0.53 | [-2.50; 1.45] | 0.457 |
| 0.3 | 0.00 | [-0.41; 0.41] | 0.998 |
| 0.7 | 0.01 | [-0.39; 0.41] | 0.919 |

**Table S2** Sensitivity analyses of pooled (standardized) mean differences (SMDs, MDs) under pre–post correlation assumptions (r = 0.3 and r = 0.7). UPDRS, Tremor, SICI. Tremor denotes a pooled tremor-severity outcome synthesized using SMD from heterogeneous endpoints, including clinician-rated tremor subscores (UPDRS-derived) and instrumental measures (accelerometry/MEG-derived tremor metrics).
